# Understanding COVID-19 testing pathways in English care homes to identify the role of point-of-care testing: an interview-based process mapping study

**DOI:** 10.1101/2020.11.02.20224550

**Authors:** Massimo Micocci, Adam L Gordon, A. Joy Allen, Timothy Hicks, Patrick Kierkegaard, Anna McLister, Simon Walne, Peter Buckle, on behalf of the CONDOR study team

**Affiliations:** NIHR London In Vitro Diagnostics Co-operative, London, UK; Division of Medical Sciences and Graduate Entry Medicine, University of Nottingham, UK; NIHR Applied Research Collaboration East Midlands (ARC-EM), Nottingham UK; NIHR Newcastle In Vitro Diagnostics Co-operative, Newcastle University, UK; Newcastle upon Tyne Hospitals NHS Foundation Trust, UK

**Keywords:** Homes for the Aged, COVID-19, Point-of-care testing, diagnosis

## Abstract

**Introduction:** Care home residents are at high risk of dying from COVID-19. Regular testing producing rapid and reliable results is important in this population because infections spread quickly and presentations are often atypical or asymptomatic. This study evaluated current testing pathways in care homes to explore the role of point-of-care tests (POCTs).

**Methods:** Ten staff from eight care homes, purposively sampled to reflect care organisational attributes that influence outbreak severity, underwent a semi-structured remote videoconference interview. Transcripts were analysed using process mapping tools and framework analysis focussing on perceptions about, gaps within, and needs arising from, current pathways.

**Results:** Four main steps were identified in testing: infection prevention, preparatory steps, swabbing procedure, and management of residents. Infection prevention was particularly challenging for mobile residents with cognitive impairment. Swabbing and preparatory steps were resource-intensive, requiring additional staff resource. Swabbing required flexibility and staff who were familiar to the resident. Frequent approaches to residents were needed to ensure they would participate at a suitable time. After-test management varied between sites. Several homes reported deviating from government guidance to take more cautious approaches, which they perceived to be more robust.

**Conclusion:** Swab-based testing is organisationally complex and resource-intensive in care homes. It needs to be flexible to meet the needs of residents and provide care homes with rapid information to support care decisions. POCT could help address gaps but the complexity of the setting means that each technology must be evaluated in context before widespread adoption in care homes.

**Key-points:**

1. Testing for COVID-19 in care homes is complex and requires reconfiguration of staffing and environment.
2. Isolation and testing procedures are challenged when providing person-centred care to people with dementia.
3. Point-of-care testing results could give care homes greater flexibility to test in person-centred ways.
4. There was evidence that care home staff interpret testing guidance, rather than follow it verbatim.
5. Each POCT must be evaluated in the context of care homes to understand its effect on care home processes.

## Introduction

Around 430,000 people in England and Wales live in care homes [1]. The majority of care home residents are older, affected by prevalent multimorbidity, activity limitation and cognitive impairment [2]. In the first six months of 2020, there were 29,393 excess deaths in care homes in England and Wales, with 19,394 attributed to COVID-19 [3].

Once a COVID-19 outbreak starts, the virus can spread rapidly through a care home. Presentations in residents are often atypical or asymptomatic. A study of 394 residents of four London care homes conducted in April 2020 [4], found 33% of residents with COVID-19 were asymptomatic. A further 31% had symptoms commonly seen in acute frailty syndromes including delirium, postural instability and diarrhoea. The high prevalence of asymptomatic or atypical presentations means that testing for the presence of SARS-CoV-2 in respiratory secretions is central to COVID-19 management. Several testing strategies have been used for residents and staff during the pandemic: an initial strategy of testing symptomatic residents only [5], progressed to a programme of 28-day and 7-day regular surveillance testing of residents and staff respectively [6]. Testing uses nasopharyngeal swabs which are sent for laboratory-based Reverse Transcriptase Polymerase Chain Reaction (RT-PCR). Frequent changes to testing protocols in the first part of the pandemic led to uncertainty as care homes had to readapt swabbing procedures and infection prevention measures multiple times, whilst the demands placed on the testing system by the rapid escalation of testing have led to delays with test results that compromise care homes’ ability to deliver effective care.

Rapid diagnostic point of care testing (POCT) could potentially address these challenges. However, little is known about the most effective way to implement these tests into existing procedures and COVID-19 management in the care home setting.

In this paper, we describe research undertaken to understand how testing strategies have been implemented in care homes, how these strategies influence the testing and management of residents and the degree of readiness in care homes for implementation of POCT.

## Methods

Between July and August 2020, care home staff members were contacted through a national online COVID-19 peer-support group for care home managers and staff [7]. Purposive sampling was used to ensure the opinions elicited were representative of a range of organisational factors (care home size, residential/nursing, independent operator/chain) that have been shown to influence the severity of outbreaks during the pandemic [8].

After an initial email contact, potential participants were asked to sign a consent form. Each participant was interviewed by an expert in Human Factors (MM) and notes were taken by an expert in process modelling (TH). They were interviewed individually and remotely, using videoconferencing tools (Zoom, Zoom Video Communications, Inc.). Interviews were semi-structured, lasted 30-60 minutes, were recorded with permission and transcribed (not verbatim). The interview schedule covered staff training and experience, and current COVID-19 testing processes. Interview transcripts were analysed using process mapping tools to describe and visualise clinical pathways [9]. Framework analysis was used to analyse transcripts against initial themes of perceptions about gaps within and needs arising from, current pathways. Codes were added to transcripts in Microsoft Excel and categories; themes were refined through a repeated analysis conducted by a second analyst (PK) – Appendix II. Participants were contacted by the research team upon completion of preliminary analysis to verify findings. No discrepancies between findings and feedback from participants were reported.

## Results

Ten staff members from eight care homes - with more than five years’ experience in the sector - accepted to take part in the study - Appendix I.

Testing for COVID-19 requires each care home to order testing kits, to swab residents, to upload each test barcodes to a dedicated portal, and to ship samples to the laboratory via pre-arranged courier. Four main steps were identified in the COVID testing and management pathway illustrated as a process in Figure 1. Table 1 summarises relevant stakeholders, guidance, resources, gaps in the pathway, needs, and opportunities for POCT in care homes.

**Table 1.**
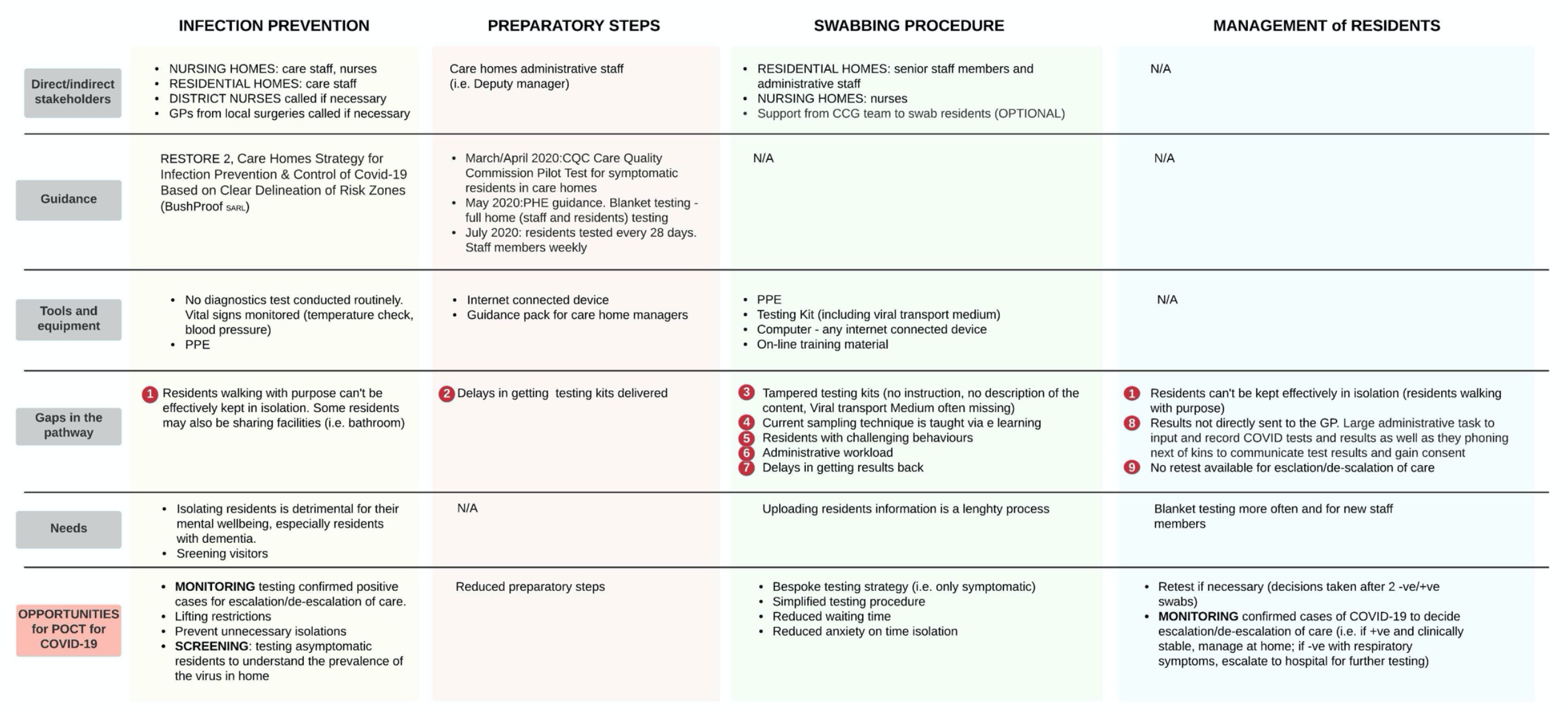
Summary of relevant stakeholders, guidance, resources, gaps in the pathway, needs and opportunities for POCT

**Fig. 1.**
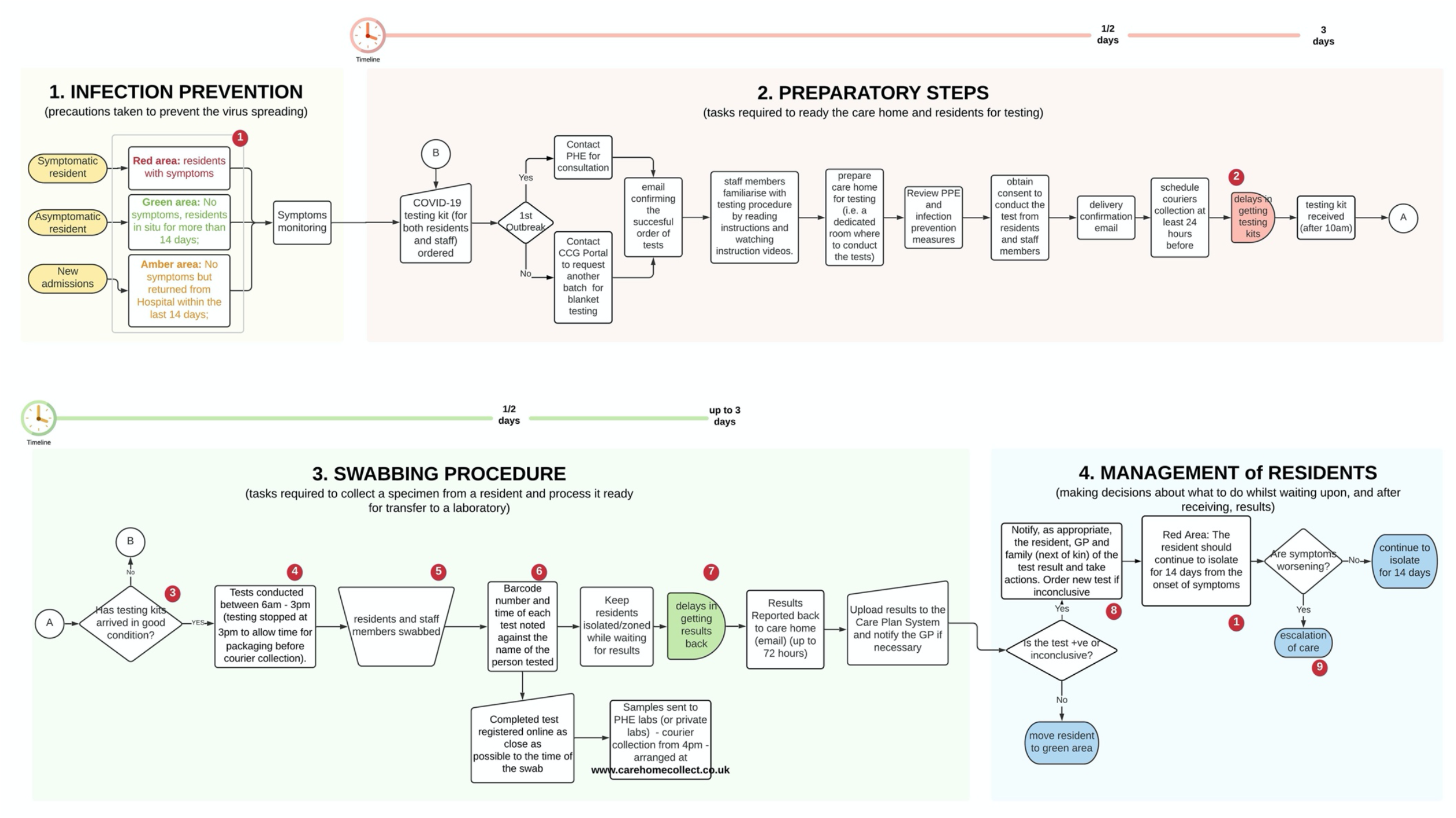
Overall swabbing and management process of resident in care homes.

The four main steps were:

1. **Infection prevention**: the allocation of residents to dedicated containment zones to prevent infections has become widespread in care homes during the pandemic [10]. Effective zoning depends upon recognising residents who are COVID-19 positive and moving them to a “red” area. These are separate from “green” areas, where COVID-19 negative residents receive care. A major challenge was supporting residents with dementia and those who ‘walk with purpose’ or ‘wander’ to understand and engage with infection prevention measures.
2. **Preparatory steps**: sequential steps are mandatory to prepare care homes for swabbing (Fig. 1). National guidance suggested two staff members should be involved – one to swab residents and one to record registration information. This had implications for staffing resource and rostering. A significant and persisting challenge was the need to do routine screening tests –weekly for staff and monthly for residents – alongside ad hoc testing for symptomatic residents. This was easier when the incidence of COVID-19 was low but became more challenging, from an organisational perspective, as incidence increased.
3. **Swabbing procedure**: staff recognised that testing was daunting for residents, particularly those with dementia. Attention was given to ensuring that staff familiar to each resident were involved in swabbing. Flexibility was required, with staff often returning to residents more than once to test at a time which was acceptable, with implications for staff time. Staff were required to register the swab, once taken, by entering data on the online portal, a process considered cumbersome and time-consuming. These complex considerations had to be addressed under time pressure because the staff were given 72-hours to complete each test from kit delivery.
4. **Management of residents**: symptomatic residents were usually asked to remain in their rooms until a test result was available. This was not always possible with residents walking with purpose. Asymptomatic residents undergoing routine testing were not restricted in their movements. Test results were returned by email, then had to be communicated to residents, families and General Practitioners, and entered in care records. Some care homes interpreted government recommendations [11] differently: several respondents considered that retesting positive residents after quarantine would provide reassurance they were no longer infective. Others suggested that repeat testing should be done in residents where there was a high suspicion of COVID-19, therefore in isolation, when a negative test returned.

## Discussion

These findings illustrate the complexity of the processes in testing care home residents for COVID-19. Infection prevention and testing processes are challenged by the individual needs of residents with dementia. Routine testing has staffing and organisational implications. Existing test registration systems place an administrative burden on staff. Current training materials are generic, with no face-to-face training and without considering complex organisational issues around testing. Also, nasopharyngeal/oropharyngeal swabs are unpleasant and alternative, less invasive processes (e.g. saliva testing) should be considered.

The variation in how guidance was interpreted by care homes, with consequential discrepancies in management approaches, illustrates the need for caution. Care home managers require a robust testing strategy to constantly monitor residents and to safeguard vulnerable people. Guidelines have not been adapted to the care home setting and, as a result, care home managers interpret them according to the needs of their unique care environment. Also, interpreting diagnostic test results requires nuanced consideration of sensitivity and specificity and how these are influenced by the prevalence of COVID-19 [12-14].

We identified several ways in which POCTs could help. They could reduce the administrative burden associated with requesting and registering tests and provide staff with greater flexibility to accommodate the needs of residents with dementia. The rapid results provided by POCTs could be beneficial in testing visitors and to allow more efficient use of zoning to save residents from prolonged and unnecessary isolation. POCT would better inform decisions about hospital admission. However, conducting a diagnostic test requires face-to-face training with professionals trained in competency assessment, test interpretation, and risk assessment around testing kits and the environment in which they will be used. Also, consideration needs to be given to how to help care homes staff interpret and respond to POCT results without introducing unacceptable variation in practice and what the role of clinicians in this process would be.

Given the vulnerability of care home residents to COVID-19 and the scale of the outbreak in the first wave, our findings have great importance to inform future management of the pandemic in care homes. There are examples of POCTs being deployed in a wide range of settings during the pandemic – such as airports [15] and universities [16, 17] – without considering context-specific issues that might influence utility. The evidence presented here suggests that such an approach will not work in care homes due to the complexity of the processes involved and context-specific evaluation should be mandatory.

The main limitation of this study is the small number of interviews. The findings cannot be regarded as representative of all care homes. They are, however, sufficient to understand and illustrate the complexity of the testing pathway in care homes as a basis for future POCT research in this setting.

## Data Availability

The data that support the findings of this study are available on request from the corresponding author, [MM]. The data are not publicly available due to their containing information that could compromise the privacy of research participants.

## Acknowledgements

The authors would like to thank care home managers and staff members who took part in the study and members of the CONDOR platform for their comments: Prof Richard Body, Prof Gail Hayward, Prof Daniel Lasserson, Dr Brian Nicholson, Dr David Ashley Price, Dr Charles Reynard, Ms Val Tate, Prof Mark Wilcox.

## Conflicts of interests

None to declare

## Authorship statement

The CONDOR study team comprises partners from Manchester University NHS Foundation Trust’s Diagnostic and Technology Accelerator (DiTA), AHSN North East and North Cumbria (NENC), UK National Measurement Laboratory, University of Manchester, University of Nottingham, University of Oxford, Yorkshire and Humber AHSN, and NIHR MedTech and In Vitro Diagnostics Co-operatives (MICs) based in Oxford, Leeds, London and Newcastle.

## Ethical approval

This project was approved as a Service Evaluation by Imperial College Healthcare NHS Trust (ICHNT) – registration no. 471.

### Funding

This work was supported by UK Research and Innovation (UKRI), Asthma UK and the British Lung Foundation, as a part of the CONDOR study. MM, PK, AML, SW, and PB are supported by the NIHR London In Vitro Diagnostics Co-operative; ALG is funded in part by the NIHR Applied Research Collaboration-East Midlands (ARC-EM); AJA and TH are supported by the NIHR Newcastle In Vitro Diagnostics Co-operative. The views expressed are those of the authors and not necessarily those of the funders, the NHS, the NIHR or the Department of Health and Social Care.

## Appendix I

**Table 2.** Description of participants taking part in the study

| Interviewee's role | Identifier | Area | Nursing Home | Residential Home | TOT number of beds |
| --- | --- | --- | --- | --- | --- |
| Owner | CH1 | Nottinghamshire |  | x | 25+ |
| Care Home Manager | CH2 | Nottinghamshire | x |  | 50+ |
| Care Home Manager | CH3 | Derbyshire | x |  | 50+ |
| Nurse | CH4 |  |  |  |  |
| Managing Director | CH5 | Nottinghamshire |  | x | 140+ |
| Project Improvement Officer | CH6 |  |  |  |  |
| Care Home Manager | CH7 | Nottinghamshire | x |  | 50+ |
| Operations Director | CH8 | Yorkshire | x | x | 100+ and 50+ apartments |
| General Manager | CH9 | Yorkshire |  | x | 40+ and 40+ apartments |
| Head of Operations | CH10 | Yorkshire and Oxfordshire | x | x | 400+ |

## Appendix II

**Table 3.** Description of four themes identified with illustrative quotes

| Theme | Description | Illustrative quotes |
| --- | --- | --- |
| Infection prevention measure | Consisting of precautions taken to prevent the virus from spreading in care homes, including new admissions and visitors' policy, and relocation of suspected cases in dedicated areas | "[following the] BushProof guidance, we have converted the whole place in cohosting zones and move people around. Red amber and green areas where created." (CH2)<br>"Extra hygiene measures and cleaning precautions taken for each room; social areas were rearranged to preserve social distancing, group activities suspended and visits from healthcare professionals (i.e. physiotherapists, speech therapists, district nurses, dieticians, GPs) were temporarily moved to a remote modality. No visitors were allowed since March; however, we have put with PVC walls [between residents and visitors]" (CH3).<br>"We have designated a separate staff room is to keep red team separated and to store uniforms." (CH8) |
| Preparatory steps | Lists of required tasks to prepare the care home, the personnel and residents to get ready for the test. These tasks include ordering testing kits, arranging courier, setting up the required environment to test residents (i.e. a dedicated room as opposed to in-room testing) and to train personnel | "nurses and senior staff members (managerial level) did the test. Video training conducted. More efficient [than symptomatic test], results come back within 24 hours. The manager was inserting patient details into the portal. Not a difficult process, but time consuming. Courier arranged from 4pm till night. It worked quite well (in early days they did not turn up)". (CH2)<br>"We struggled to get access to tests at the beginning (COVID was already in home, 3 suspected residents)" CH5)<br>For nursing staff, managing the procedure hasn't been an issue: "Training video was factual, succinct, straight to the point, easy to follow". Swabbing procedures "are not necessarily above the abilities of non-trained staff, unless training is followed properly and conducted precisely". (CH4) |
| Swabbing procedure | Tasks required to collect a specimen from a resident and process it ready for transfer to a laboratory | "The registration portal is a link after a link, it can be a nightmare" (CH3)<br>"The Digital Portal is very easy and straightforward [however] it is very time-consuming uploading test results and then once back, re-uploading them to the care plan system and communicate results to the system". (CH5) |
|  |  | *"Very difficult testing dementia residents; we adopt distraction techniques, gamification techniques". (CH2)*<br>*"We had to employ a great deal of reassurance for dementia residents; It is very important that tests are conducted by familiar faces. They trust individuals in their own spaces [...] No external individuals can pretend residents are concordant with the testing timing". CH4*<br>*"Out of the pandemic context, we would be told not to do something unless trained to do so - are you competent to do these things?" (CH9)* |
| Management of residents | Consisting of measures, treatment and isolation decisions taken upon test results and whilst waiting on test results | The isolation has a very bad effect on residents. In some cases, *"isolation is worse than the virus..."*. (CH7)<br>Treating each resident as suspected (following the guidance) is not ideal: *"If a negative resident is considered a suspected case, he may end-up isolated with positive residents"*. (CH10) |

## Notes

### Competing Interest Statement

The authors have declared no competing interest.

### Author Declarations

This project was approved as a Service Evaluation by Imperial College Healthcare NHS Trust (ICHNT), registration no. 471

